# Human milk feeding, fortification initiation, and clinical outcomes in neonates with critical congenital heart disease: A multi-institutional study

**DOI:** 10.64898/2026.08.20.26360934

**Authors:** Kristin M. Elgersma, Brian F. Joy, Zuofu Huang, Monique R. Radman, Kimberly I. Mills, Jennifer E. Schramm, Joshua H. Wong, Meghan M. Chlebowski, Asaad G. Beshish, Raya Safa, Dana Mueller, Brittany L. Shutes, Chetna Pande, Jamie Furlong-Dillard, Sukumar Suguna Narasimhulu, Andrea Beach, Stephanie A. Goldstein, Christine M. Riley, Reshma Reddy, Jordan Schneider, Nimrod Goldshtrom, Grace Liao, Ahmed Asfari, Karan B. Karki, Robert Marcel T Huibonhoa, Christopher W. Mastropietro, Katherine Cashen

## Abstract

**Background:** Neonates with critical congenital heart disease (CCHD) are vulnerable to feeding-related complications including necrotizing enterocolitis (NEC). Human milk and direct breastfeeding (BF) may offer protection, but multisite evidence is limited. We aimed to determine relationships between the proportion of human milk received (ie, human milk percentage) or BF frequency during the neonatal period and NEC, sepsis, infectious complications, or length of stay (LOS). We also determined whether bovine-derived fortification or formula initiation was associated with NEC.

**Methods:** This retrospective study included neonates from 25 US pediatric centers who underwent surgery with cardiopulmonary bypass. Outcomes were NEC (modified Bell’s Stages II–III), sepsis, infection, and LOS. Disease risk score case-control matching and energy balancing weighted regression balanced multiple relevant covariates.

**Results:** Among 822 neonates, the percentage of human milk received during the neonatal period was not associated with NEC, sepsis or infection. Initiation of fortification or formula was associated with 3-fold higher odds of developing NEC within 5 days (OR:3.10, 95%CI:1.10–8.12, p=0.025). In energy balancing weighted regression models, higher neonatal human milk percentage and more frequent BF were strongly associated with shorter LOS: 100% versus 0% human milk with 9.33 days shorter (4.47–14.19, p<0.001); each additional BF session with 0.48 days shorter (0.31–0.65, p<0.001).

**Conclusions:** In this multisite cohort, fortification or formula initiation was associated with increased odds of NEC; and neonatal human milk percentage and BF with shorter LOS. Given limited evidence to guide practice, caution in introducing bovine-derived formula for high-risk infants with CCHD may be warranted.

**Clinical Perspective:** *What Is New?:*

- This study including 822 neonates with critical congenital heart disease from 25 US pediatric cardiac centers identified a relationship between bovine-derived fortification or formula initiation and increased likelihood of developing necrotizing enterocolitis (modified Bell’s stage II–III) within 5 days.
- A higher proportion of human milk and more frequent direct breastfeeding during the neonatal period were strongly associated with a shorter hospital length of stay, even when accounting for more than 20 relevant clinical variables.

*What Are the Clinical Implications?:*

- Clinical support of human milk and direct breastfeeding is important for infants born with critical congenital heart disease. Based on our findings, caution in introducing bovine-derived fortification or formula for high-risk infants with critical congenital heart disease may be warranted.

## Introduction

While critical congenital heart disease (CCHD) remains a leading cause of death for children globally and in the United States (US),^1^ improvements in care over the past several decades have substantially reduced mortality, particularly during infancy.^2^ Clinical complications and prolonged hospital length of stay (LOS), however, are still common and associated with worse long-term outcomes,^3^ contributing to an average lifetime cost of over $2 million per patient.^4^ Much of this cost is concentrated in early childhood, a time where feeding challenges play a key role.^5^ For example, necrotizing enterocolitis (NEC), a severe gastrointestinal complication, occurs most commonly between 22 and 25 days of age among patients with CCHD^6^ and has been associated with more than $350,000 greater hospital costs and 36 days longer hospital LOS.^7^ NEC and other feeding challenges are highly stressful for families, who often cite feeding as their number one concern – more so than their child’s cardiac condition.^8^ Unfortunately, evidence to guide CCHD-specific nutrition and feeding regimens remains limited, leading to substantial practice variation among providers and across centers.^9–11^

Human milk and direct breastfeeding (BF) are the optimal, biologically-normative nutrition for all newborns, including those who require hospitalization.^12,13^ Most evidence linking human milk to improved short- and long-term outcomes in hospitalized infants originates from preterm populations, where multiple systematic reviews and meta-analyses have demonstrated a convincing reduction in NEC risk.^14–16^ A 2024 Cochrane review of studies including very preterm and low-birth weight infants found that supplementation with donor human milk, compared to commercial formula, reduced NEC by nearly 50% (95%CI: 24%–63%),^14^ and another meta-analysis suggests that this effect may be dose-dependent.^15^ Although the etiology of cardiac NEC in CCHD may differ from preterm NEC (eg, greater contribution of splanchnic hypoxia),^17^ emerging evidence similarly links higher human milk intake with reduced NEC in CCHD populations.^18–21^ Recent studies have reported lower preoperative NEC among infants fed exclusive, unfortified human milk;^18^ lower pre- and postoperative NEC for infants with single ventricle physiology who have high human milk exposure;^20^ lower NEC rates with exclusive human milk diets including human milk-derived fortification,^19^ and increased postoperative NEC for infants who were formula fed on their last preoperative day.^21^ Beyond NEC, human milk and BF may decrease sepsis and infection rates and reduce LOS for infants with CCHD.^20,22,23^

The existing CCHD literature is limited by a lack of detailed, daily nutrition data; sample sizes that are underpowered to detect modified Bell’s stage II–III NEC (a rarer but more rigorously defined outcome);^24^ unclear alignment of outcome timing with nutritional exposures; and insufficient data on fortification practices.^23^ The current standard for high-quality neonatal human milk research involves measurement of daily nutritional exposures.^25–29^ To our knowledge, multisite research utilizing daily nutrition data for newborns with CCHD is limited to one study,^19^ which was primarily focused on the effect of a human-milk derived fortifier on growth outcomes. Furthermore, few CCHD studies have investigated relationships between BF and clinical outcomes, despite distinct potential microbiome, cardiorespiratory, and neurodevelopmental benefits related to direct BF.^30–32^ To address these limitations, we aimed to estimate the effects of the proportion of human milk received (ie, human milk percentage) and BF frequency on NEC (Bell’s stage II-III), sepsis, infectious complications, and LOS in a multisite cohort including daily nutrition data. We also sought to assess whether bovine-derived fortification or formula initiation was associated with the development of NEC.

## Methods

### Study design

This retrospective study incorporated cohort and case-control approaches and was conducted across 25 United States pediatric cardiac centers participating in Collaborative Research from the Pediatric Cardiac Intensive Care Society (CoRe-PCICS; Table S1). CoRe-PCICS member institutions establish data use agreements with Riley Hospital for Children at Indiana University Health, which serves as the data coordinating center for all CoRe-PCICS studies. Ethical approval for the study was obtained through each site’s Institutional Review Board, with informed consent waived due to the retrospective nature of the deidentified data. The analytical methods used in this study can be made available to researchers outside the CoRe-PCICS collaborative if requested. The multisite data are governed under the conditions of the data use agreements between participating institutions and the study coordinating center and will not be publicly available.

### Population

This study built upon a previous CoRe-PCICS study cohort focused on hyperoxia during cardiopulmonary bypass (CPB),^33^ which included neonates born in 2021-2022 who underwent Society of Thoracic Surgeons-European Association for Cardio-Thoracic Surgery (STAT) mortality category 2-5 cardiac surgery with CPB. For the current study, participants were less than 7 days old at hospital admission to ensure consistency in early nutrition establishment and lactation support. A maximum of 50 patients were included per center, with participants chosen randomly as needed. Exclusion criteria for the initial hyperoxia cohort were corrected gestational age less than 37 weeks at surgery, preoperative cardiopulmonary resuscitation or extracorporeal membrane oxygenation (ECMO), failure to separate from CPB necessitating postoperative ECMO, non-CPB cardiac intervention prior to CPB surgery (eg, shunt placement, pulmonary artery banding), or cardiac transplantation as initial surgery. Additional exclusion criteria for this analysis included discharge before initial surgery or after 6 months corrected age, less than 7 days of enteral feeding in the neonatal period (defined as the first 28 postnatal days), and a major congenital anomaly that could impact enteral or oral feeding (eg, cleft palate). For the LOS outcome, infants were excluded if they died before discharge. Figure 1 is a flow diagram outlining the analytical cohorts.

**Figure 1.**
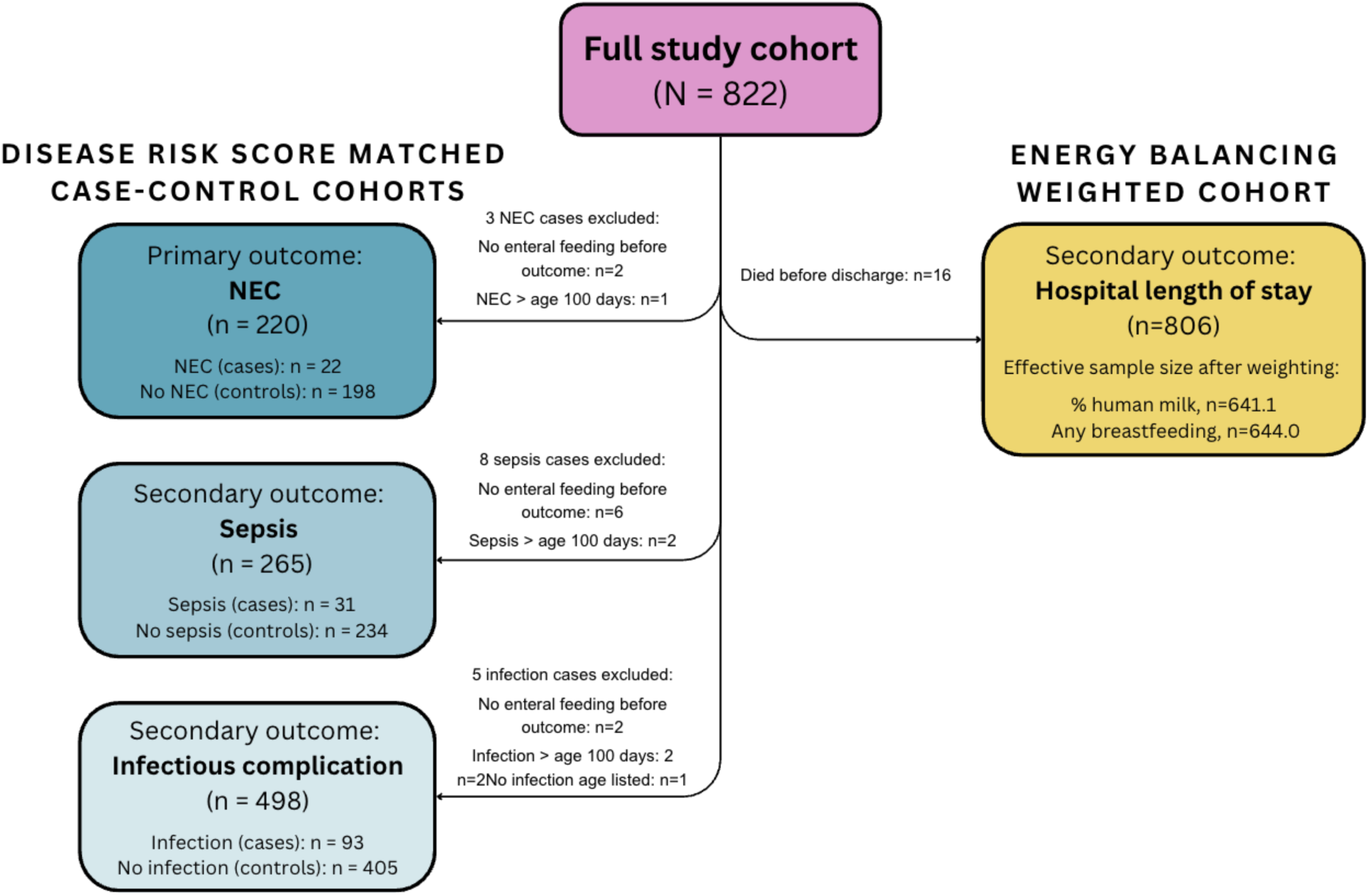
Flow diagram of all study analytical cohorts

### Data collection

Data were collected from the electronic health record (EHR) and entered into a study REDCap by site principal investigators (PIs). As most CoRe-PCICS member sites also contribute data to the well-established Pediatric Cardiac Critical Care Consortium (PC4) registry, data definitions mirrored PC4 definitions whenever possible.^34^ Daily nutrition data included volume, type (eg, maternal or donor human milk, commercial formula), route (eg, breastfeeding, bottle, feeding tube), total parenteral nutrition, fortification type and timing, and caloric density (eg, 24 kcal/oz) for the first 28 days of life, plus the 2 days before hospital discharge and before a post-neonatal NEC diagnosis, if applicable. Data were cleaned and quality checked, with site PIs contacted for clarification when necessary.

### Outcomes

Complete outcomes definitions can be found in the Detailed Methods. The primary outcome was NEC, defined per PC4 as modified Bell’s Stages II–III,^34^ in the first 100 postnatal days. Secondary outcomes were sepsis in the first 100 days; other infectious complications in the first 100 days, defined as surgical site infection, central line-associated bloodstream infection, urinary tract infection, pneumonia, and preoperative respiratory viral infection; and hospital LOS.

### Exposures

Exposures of interest were (1) human milk percentage, defined as the proportion of neonatal enteral base diet nutrition that was maternal or donor human milk (ie, not including fortification), (2) BF frequency, defined as a direct latch at the breast, and (3) bovine-derived fortification or formula initiation within 5 days before the NEC diagnosis age (see further information in the statistical analysis section). There is no standard window for evaluating factors associated with NEC. Prior studies have examined exposures ranging from 72 hours to 2 weeks before diagnosis.^35,36^ We selected a 5-day window to align with fortification/formula initiation studies in preterm infants that evaluated 4–7 day pre-NEC periods.^37,38^ This window captures acute effects while allowing for processes that evolve over several days. Fortification was defined as any nutritional substance designed to increase the energy value of the infant’s base diet, including human milk fortifier, formula added to human milk, and formula prepared at a non-standard concentration. Due to sample size, we were not able to test for differences in fortification type (eg, hydrolyzed, amino-acid based). Only one infant in the full study cohort received human milk-derived fortifier and that infant was not included in the case-control matched analysis; therefore all infants in the NEC case-control matched analysis received bovine-derived fortifier. Direct BF volume was mostly unmeasured; thus, we estimated the volume of each BF session conservatively, based on published averages for term and hospitalized infants.^39,40^ Human milk percentage was selected as a primary exposure to characterize diet composition while minimizing confounding by feeding volume. While maternal and donor human milk differ, donor human milk use was limited in our sample and stratifying by human milk type was not possible. Further details are in the Detailed Methods.

### Statistical analysis

To test the hypothesis that increased human milk percentage or increased BF are associated with reduced NEC, sepsis, and other infectious complications, we implemented a disease risk score matched case-control approach, individualized for each outcome. Infants with the outcome were included if they were enterally fed before diagnosis, to fulfill the statistical assumption of positivity (see Figure 1). Each case was matched with up to 10 matches, resulting in excellent covariate balance between the cohorts (Figure S2, Table S2). Controls were assigned the same diagnosis age as their matched case for each outcome. For example, if a patient developed NEC at age 20 days, all 10 matched controls for that patient were assigned a pseudo “NEC diagnosis age” of 20 days, and nutrition exposures (eg, % of human milk; fortification or formula within the previous 5 days) were evaluated before that pseudo NEC diagnosis age. This case-control method addressed limitations of alternative approaches and preserved a potential causal pathway by including only nutrition data that occurred before each outcome, and in the same time period for the matched controls. In each matched cohort, we used weighted logistic regression to model the binary occurrence of the outcome as a function of each exposure. Due to low BF frequency causing separation in the models, we were limited to testing a binary BF exposure for these outcomes, ie, any BF before diagnosis age versus none. Post hoc sensitivity analyses included adjustment for maximum enteral volume; extending the timeframe for matched controls to include nutrition data both 5 days before and 5 days after their pseudo diagnosis age; including only infants with a diagnosis age of 30 days or less; and adjustment for broad fortification types (standard or specialized; fortification types listed in Table S3). Further details including matching variables can be found in the Detailed Methods.

To test the hypothesis that increased human milk percentage or increased BF frequency are associated with reduced hospital LOS, we applied an energy balancing approach to make the covariate distributions of treatment and control groups as similar as possible.^41^ We included the full cohort, as all nutrition data inherently occurred prior to the outcome; thus, matching was not necessary. Energy balancing reweights each participant’s contribution to the analysis based on their likelihood of exposure given their characteristics, and we balanced 20 relevant covariates across the sample (see Detailed Methods). We used weighted linear regression to model hospital LOS as a function of human milk percentage and of BF frequency during the neonatal period. Post hoc sensitivity analyses included further adjustment for multiple clinical complications, dichotomization of exposures (eg, any BF; any preoperative BF, “high” human milk ≥90%), and subgroup analysis of infants with single ventricle physiology. There were no missing data in the full sample. For variables that were unknown or undocumented in the EHR an “Unknown” category was used and included in the matching or weighting approaches. Affected covariates were Hispanic ethnicity (n=60 unknown) and birth delivery mode (n=12 unknown). The corresponding author (KME) had full access to all the data in the study and takes responsibility for its integrity and the data analysis. Significance was set at p<0.05, and analyses were performed in R version 4.5.1.

## Results

There were 822 patients in the full study cohort, with all analytical cohorts (case matching and energy balancing weighting) drawn from this larger group. Sample characteristics can be found in Table 1. A total of 226 (27.5%) participants had single ventricle physiology, 423 (51.5%) had a STAT score of 4 or 5, the median (25%, 75%) age at surgery of 6 (4, 8) days old, and the median hospital LOS was 26 (18, 40) days. Overall, 25 (3.0%) infants were diagnosed with NEC (n=15, 6.6% of all infants with single ventricle CCHD; n=10, 1.7% of infants with non-single ventricle CCHD). A total of 39 (4.7%) had sepsis, and 98 (11.9%) experienced the composite “other infectious complication.” Patients consumed, on average, 70.1% (±36.8%) of their enteral volume as human milk during the neonatal period, of which 2.9% was donor human milk. Overall, 183 (22.3%) infants across 20 (80%) sites received donor human milk at least once, and 233 (28.3%) were directly breastfed at least once. Further details of the matched cohorts are in Table 2 (NEC), Table S4 (sepsis), and Table S5 (other infectious complications).

**Table 1.** Sample characteristics (n=822)

|  | n (%), mean (SD), or median (25%, 75%)* |
| --- | --- |
| Sex at birth |  |
| Male | 513 (62.4) |
| Female | 308 (37.5) |
| Ambiguous | 1 (0.1) |
| Race |  |
| White | 542 (65.9) |
| Black/African American | 116 (14.1) |
| Another race or multirace | 164 (20.0) |
| Ethnicity |  |
| Hispanic/Latino | 146 (17.8) |
| Not Hispanic/Latino | 616 (74.9) |
| Not documented | 60 (7.3) |
| Insurance status |  |
| Public | 424 (51.6) |
| Private, self, or other | 398 (48.4) |
| Child Opportunity Index | 51 (28, 76) |
| Prenatal CHD diagnosis | 585 (71.2) |
| Birth WAZ | -0.20 (1.05) |
| Single ventricle physiology | 226 (27.5) |
| Genetic syndrome | 141 (17.2) |
| Extracardiac abnormality | 159 (19.3) |
| Any preoperative feeding | 595 (72.4) |
| Surgery age (days) | 6 (4, 8) |
| Weight (kg) at surgery | 3.31 (0.51) |
| STAT score 4 or 5 | 423 (51.5) |
| Sternum left open | 381 (46.4) |
| Postoperative ECMO | 36 (4.4) |
| Necrotizing enterocolitis† | 25 (3.0) |
| Sepsis | 39 (4.7) |
| Other infectious complication‡ | 98 (11.9) |
| NEC diagnosis age (days) | 20 (15, 30) |
| % of volume as human milk (base diet) during the neonatal | 70.1 (36.8) |
| Exclusive human milk (base diet) during the neonatal period | 290 (35.3) |
| Any breastfeeding during the neonatal period | 233 (28.3) |
| Length of stay (days) | 26 (18, 40) |

Notes:
\*Descriptive statistics are n (%) for categorical variables, mean (SD) for continuous variables except median (25%, 75%) for Child Opportunity Index, age at surgery, NEC diagnosis age, and hospital length of stay.
†NEC is defined as modified Bell's criteria stage II–III.
‡Infectious complications are defined as surgical site infection, central line-associated blood stream infection, urinary tract infection, pneumonia, and preoperative respiratory viral infection.
Abbreviations: CHD = congenital heart disease, ECMO = extracorporeal membrane oxygenation, STAT = Society of Thoracic Surgeons-European Association for Cardio-Thoracic Surgery, WAZ = weight-for-age z-score

**Table 2.**
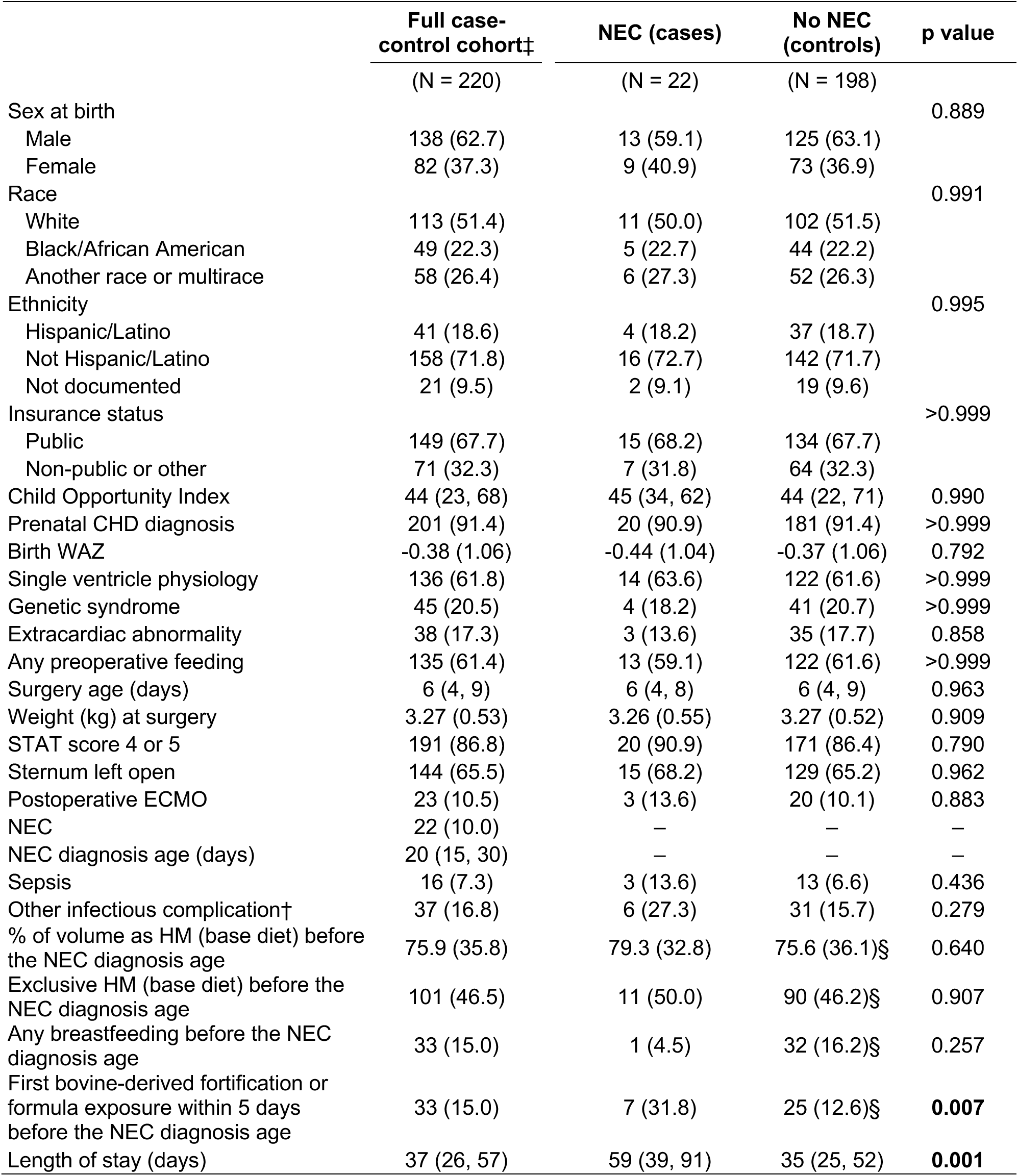
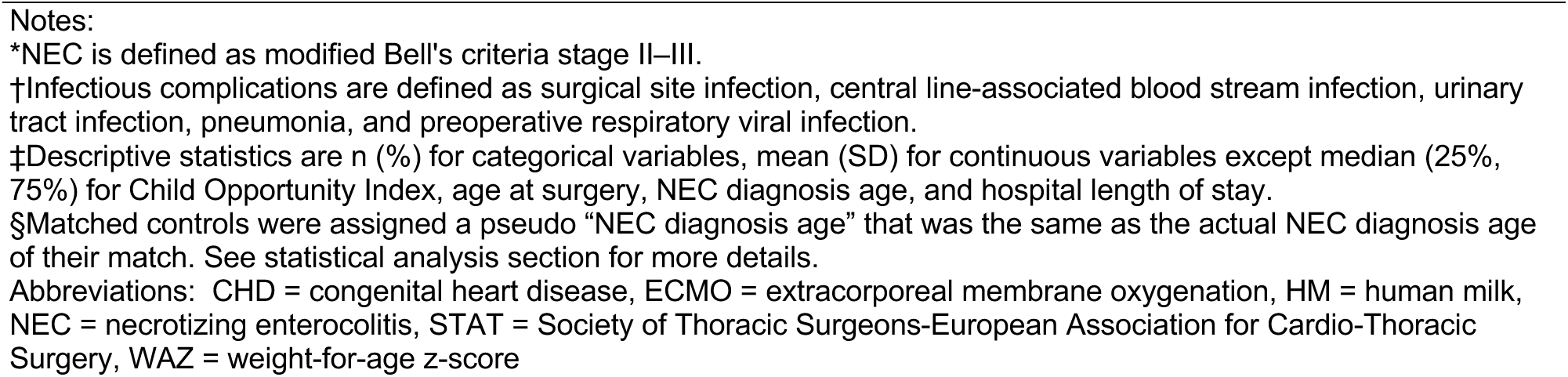
Characteristics of the disease risk score matched cohort for the necrotizing enterocolitis outcome*.

|  | Full case-control cohort‡ | NEC (cases) | No NEC (controls) | p value |
| --- | --- | --- | --- | --- |
|  | (N = 220) | (N = 22) | (N = 198) |  |
| Sex at birth |  |  |  | 0.889 |
| Male | 138 (62.7) | 13 (59.1) | 125 (63.1) |  |
| Female | 82 (37.3) | 9 (40.9) | 73 (36.9) |  |
| Race |  |  |  | 0.991 |
| White | 113 (51.4) | 11 (50.0) | 102 (51.5) |  |
| Black/African American | 49 (22.3) | 5 (22.7) | 44 (22.2) |  |
| Another race or multirace | 58 (26.4) | 6 (27.3) | 52 (26.3) |  |
| Ethnicity |  |  |  | 0.995 |
| Hispanic/Latino | 41 (18.6) | 4 (18.2) | 37 (18.7) |  |
| Not Hispanic/Latino | 158 (71.8) | 16 (72.7) | 142 (71.7) |  |
| Not documented | 21 (9.5) | 2 (9.1) | 19 (9.6) |  |
| Insurance status |  |  |  | >0.999 |
| Public | 149 (67.7) | 15 (68.2) | 134 (67.7) |  |
| Non-public or other | 71 (32.3) | 7 (31.8) | 64 (32.3) |  |
| Child Opportunity Index | 44 (23, 68) | 45 (34, 62) | 44 (22, 71) | 0.990 |
| Prenatal CHD diagnosis | 201 (91.4) | 20 (90.9) | 181 (91.4) | >0.999 |
| Birth WAZ | -0.38 (1.06) | -0.44 (1.04) | -0.37 (1.06) | 0.792 |
| Single ventricle physiology | 136 (61.8) | 14 (63.6) | 122 (61.6) | >0.999 |
| Genetic syndrome | 45 (20.5) | 4 (18.2) | 41 (20.7) | >0.999 |
| Extracardiac abnormality | 38 (17.3) | 3 (13.6) | 35 (17.7) | 0.858 |
| Any preoperative feeding | 135 (61.4) | 13 (59.1) | 122 (61.6) | >0.999 |
| Surgery age (days) | 6 (4, 9) | 6 (4, 8) | 6 (4, 9) | 0.963 |
| Weight (kg) at surgery | 3.27 (0.53) | 3.26 (0.55) | 3.27 (0.52) | 0.909 |
| STAT score 4 or 5 | 191 (86.8) | 20 (90.9) | 171 (86.4) | 0.790 |
| Sternum left open | 144 (65.5) | 15 (68.2) | 129 (65.2) | 0.962 |
| Postoperative ECMO | 23 (10.5) | 3 (13.6) | 20 (10.1) | 0.883 |
| NEC | 22 (10.0) | — | — | — |
| NEC diagnosis age (days) | 20 (15, 30) | — | — | — |
| Sepsis | 16 (7.3) | 3 (13.6) | 13 (6.6) | 0.436 |
| Other infectious complication† | 37 (16.8) | 6 (27.3) | 31 (15.7) | 0.279 |
| % of volume as HM (base diet) before the NEC diagnosis age | 75.9 (35.8) | 79.3 (32.8) | 75.6 (36.1)§ | 0.640 |
| Exclusive HM (base diet) before the NEC diagnosis age | 101 (46.5) | 11 (50.0) | 90 (46.2)§ | 0.907 |
| Any breastfeeding before the NEC diagnosis age | 33 (15.0) | 1 (4.5) | 32 (16.2)§ | 0.257 |
| First bovine-derived fortification or formula exposure within 5 days before the NEC diagnosis age | 33 (15.0) | 7 (31.8) | 25 (12.6)§ | **0.007** |
| Length of stay (days) | 37 (26, 57) | 59 (39, 91) | 35 (25, 52) | **0.001** |

\*NEC is defined as modified Bell's criteria stage II–III.
†Infectious complications are defined as surgical site infection, central line-associated blood stream infection, urinary tract infection, pneumonia, and preoperative respiratory viral infection.
‡Descriptive statistics are n (%) for categorical variables, mean (SD) for continuous variables except median (25%, 75%) for Child Opportunity Index, age at surgery, NEC diagnosis age, and hospital length of stay.
§Matched controls were assigned a pseudo “NEC diagnosis age” that was the same as the actual NEC diagnosis age of their match. See statistical analysis section for more details.
Abbreviations: CHD = congenital heart disease, ECMO = extracorporeal membrane oxygenation, HM = human milk, NEC = necrotizing enterocolitis, STAT = Society of Thoracic Surgeons-European Association for Cardio-Thoracic Surgery, WAZ = weight-for-age z-score

### Human milk percentage, BF, and NEC, sepsis, or other infectious complications

In the case-matched cohorts, the percentage of human milk consumed before the diagnosis age was not associated with diagnosis of NEC, sepsis, or other infectious complications (Table 3). Similarly, although the estimated odds of each outcome were lower for infants with any BF, none of these associations reached statistical significance (Table 3). Overall, the number of breastfed patients was low, particularly in the NEC, sepsis, and infection groups. Only 1 (4.5%) infant with NEC had any pre-diagnosis BF, compared to 24 (12.1%) in the matched controls; only 1 (3.2%) infant with sepsis was breastfed, compared to 35 (15%) controls; and 13 (14.3%) infants with an infectious complication were breastfed, compared to 79 (19.4%) controls.

**Table 3.** Weighted regression models of human milk percentage or any breastfeeding and NEC, sepsis, or other infectious complication*.

| **Outcome: NEC** | **OR** | **95% CI** | **p value** |
| --- | --- | --- | --- |
| % of enteral volume as human milk† | 1.35 | (0.40-5.90) | 0.656 |
| Any direct breastfeeding | 0.36 | (0.02-1.86) | 0.330 |
| **Outcome: Sepsis** |  |  |  |
| % of enteral volume as human milk | 0.58 | (0.21-1.71) | 0.297 |
| Any direct breastfeeding | 0.21 | (0.01-1.05) | 0.135 |
| **Outcome: Other infectious complication‡** |  |  |  |
| % of enteral volume as human milk | 1.53 | (0.78-3.15) | 0.230 |
| Any direct breastfeeding | 0.74 | (0.37-1.35) | 0.326 |

Note:
\*Variables accounted for in all analyses by the disease risk score matching process include birth weight-for-age z-score (WAZ), prenatal CCHD diagnosis, infant delivery mode, infant race, Hispanic ethnicity, insurance type, Child Opportunity Index 2.0 score, single ventricle physiology, aortic obstruction, major genetic syndrome, extracardiac anomaly, preoperative factors including mechanical circulatory support, shock, or neurological deficit, highest level of preoperative respiratory support, preoperative enteral feeding, prostaglandin E1 days, Society of Thoracic Surgeons–European Association for Cardio-Thoracic Surgery (STAT) mortality category, age at surgery, weight at surgery, sternum left open postoperatively, postoperative extracorporeal membrane oxygenation (ECMO).
†All nutrition calculated before the outcome diagnosis age.
‡Infectious complications were defined as surgical site infection, central line-associated blood stream infection, urinary tract infection, pneumonia, and preoperative respiratory viral infection.
Abbreviations: BF = breastfeeding, CI = confidence interval, NEC = necrotizing enterocolitis, OR = odds ratio

### Fortification or formula initiation and NEC

To investigate the relationship between fortification or formula initiation and NEC, we first visualized the nutrition patterns for each NEC case. As shown in Figure 2, while neonates with NEC received primarily human milk before diagnosis, bovine-derived fortification or formula was started a median 5 days prior to NEC diagnosis. A total of 7 (31.8%) patients with NEC had an initial exposure to fortification or formula within 5 days of diagnosis (Table 2).

**Figure 2.**
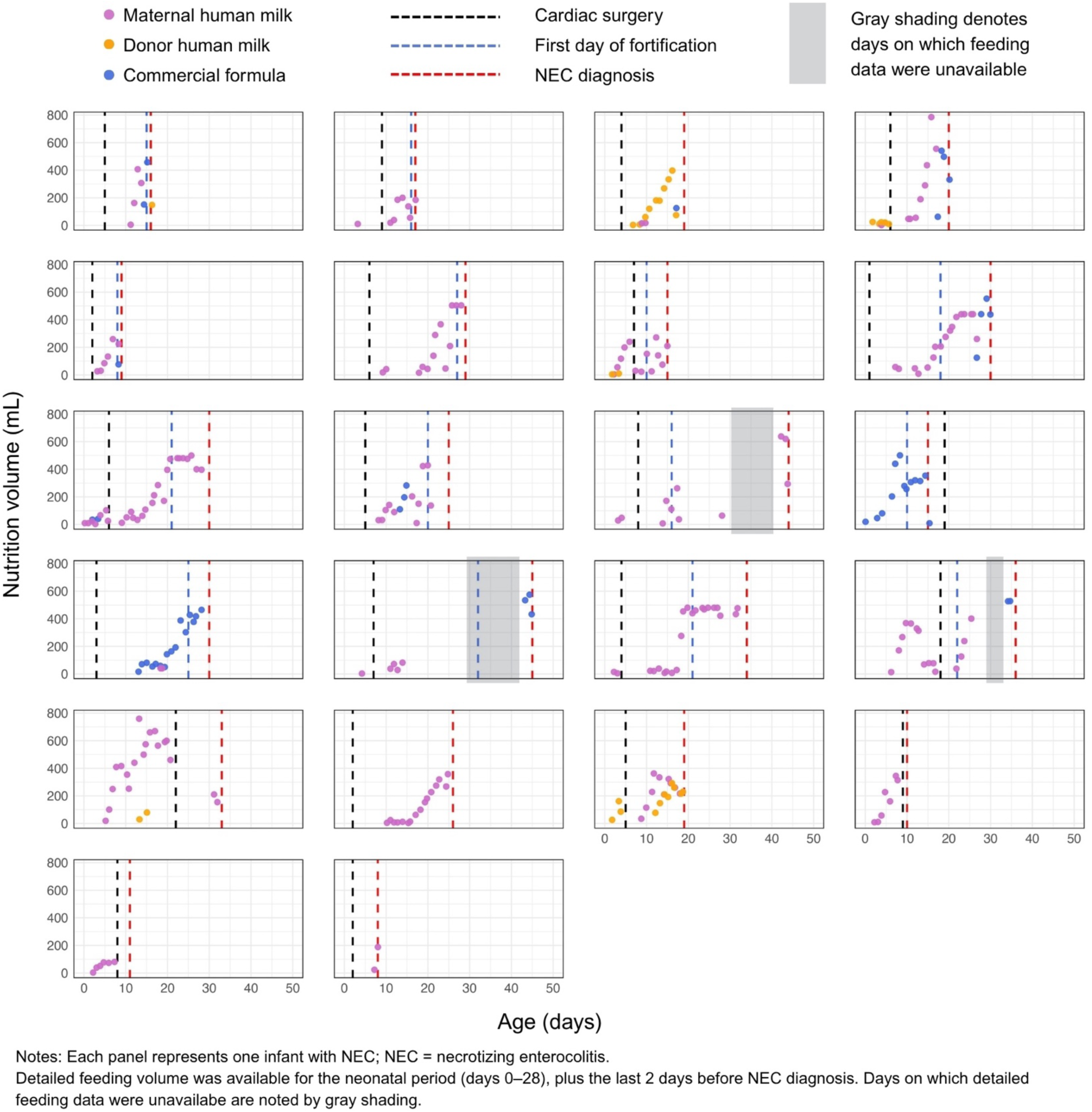
Nutrition patterns suggest a relationship between formula or fortification initiation and subsequent NEC diagnosis for infants with CCHD (n=22, age 7–45 days at diagnosis)

In disease risk score matched models (Table 4), fortification or formula initiation within 5 days before the NEC diagnosis age was associated with 3-fold higher odds of NEC (OR:3.10, 95%CI:1.10-8.12, p=0.025). This result was consistent in sensitivity analyses adjusting for pre-diagnosis maximum enteral volume (mean 133 mL/kg/d for NEC cases versus 127 mL/kg/d for controls, p=0.635); extending the timeframe to include 5 days both before and after the NEC diagnosis age; limiting diagnosis age to less than 30 days to include only neonatal nutrition data; and considering fortification or formula type (Table 4). We considered whether the fortification concentration could have played a role; however, only 27 patients (12.3%) received more than 24 kcal/oz before the NEC diagnosis age (n=2, 9% NEC cases, n=25, 13% matched controls, p=0.632). In the subset of patients who were included in the previous CoRe-PCICS hyperoxia study^33^ and for whom detailed operative data had been collected (n=192, 87%), we compared the average CPB time, cross-clamp time, and vasoactive-ventilation-renal (VVR) score^42^ 12 hours after cardiac ICU admission between the NEC cases and controls, and found no significant differences: CPB mean 160 minutes (NEC) vs. 153 minutes (controls), p=0.912; cross clamp 75 minutes (NEC) vs. 71 minutes (controls), p=0.792; VVR score 28 (NEC) vs 26 (controls), p=0.394.

**Table 4.** Weighted regression models of initial bovine-derived formula or fortification exposure within 5 days of NEC diagnosis*.

| **Outcome: NEC** | **OR** | **95% CI** | **p value** |
| --- | --- | --- | --- |
| First formula or fortification exposure within 5 days before NEC diagnosis age† | 3.10 | (1.10-8.12) | **0.025** |
| *Sensitivity analyses* |  |  |  |
| Time window expanded to within 5 days before or 5 days after NEC age | 2.74 | (1.03-7.29) | **0.044** |
| Limited to NEC cases < age 30 days | 3.43 | (1.20-9.85) | **0.022** |
| Further adjusted for maximum enteral volume (mL/kg/day) before NEC diagnosis age‡ | 2.99 | (1.108.05) | **0.031** |
| Further adjusted for fortification type (standard or specialized) §\|\| | 2.90 | (1.05-7.99) | **0.040** |

Notes:
\*NEC is defined as modified Bell's criteria stage II–III.
†Variables accounted for in all analyses by the disease risk score matching process include birth weight-for-age z-score (WAZ), prenatal CCHD diagnosis, infant delivery mode, infant race, Hispanic ethnicity, insurance type, Child Opportunity Index 2.0 score, single ventricle physiology, aortic obstruction, major genetic syndrome, extracardiac anomaly, preoperative factors including mechanical circulatory support, shock, or neurological deficit, highest level of preoperative respiratory support, preoperative enteral feeding, prostaglandin E1 days, Society of Thoracic Surgeons–European Association for Cardio-Thoracic Surgery (STAT) mortality category, age at surgery, weight at surgery, sternum left open postoperatively, postoperative extracorporeal membrane oxygenation (ECMO).
‡Estimate for maximum volume (mL/kg/day) as a covariate for adjustment in the model is 1.00 (0.99–1.01), $p=0.673$ .
§Specialized formula is defined as hydrolyzed, partially hydrolyzed, amino acid-based, or fat-modified. No infants received human milk-based fortifier (e.g., Prolacta).
||The estimate for standard fortification, compared to no fortification, as a covariate for adjustment in the model is 1.35 (0.47–3.86), $p=0.572$ . The estimate for specialized fortification, compared to no fortification, as a covariate for adjustment in the model is 0.81 (0.25–2.66), $p=0.737$ .
Abbreviations: CI = confidence interval, NEC = necrotizing enterocolitis, OR = odds ratio

### Human milk percentage, BF frequency, and hospital LOS

In weighted regression models using energy balancing weights, both human milk percentage and direct BF during the neonatal period were strongly associated with shorter hospital LOS (Table 5). Neonates with 100% human milk, compared to 0%, had an estimated 9.33 days shorter LOS (95%CI:4.47–14.19 days shorter, p<0.001), which translates to an average of nearly 1 day shorter LOS for every 10% increase in human milk. In sensitivity analyses, infants with high human milk percentage (90% or more) had an average 6.80 days shorter LOS (p<0.001).

**Table 5.** Energy balancing weighted models* of associations between human milk or direct breastfeeding during the neonatal period and hospital length of stay (n=758)

| **Outcome: Length of stay (days)** | **Estimate ($\beta$)** | **SE** | **95% CI** | **p value** |
| --- | --- | --- | --- | --- |
| *Exposure: Human milk percentage* |  |  |  |  |
| % of volume as human milk | -9.33 | 2.48 | (-14.19, -4.47) | **<0.001** |
| High human milk ($\geq 90\%$) | -6.80 | 1.86 | (-10.43, -3.16) | **<0.001** |
| Sensitivity analysis: further adjusted for clinical complications*† |  |  |  |  |
| % of volume as human milk | -5.74 | 2.19 | (-10.03, -1.45) | **0.009** |
| High human milk ($\geq 90\%$) | -5.41 | 1.53 | (-8.41, -2.41) | **<0.001** |
| *Exposure: Direct breastfeeding* |  |  |  |  |
| Number of breastfeeding sessions | -0.48 | 0.09 | (-0.65, -0.31) | **<0.001** |
| Any breastfeeding | -8.65 | 2.45 | (-13.46, -3.85) | **<0.001** |
| Any preoperative breastfeeding‡ | -6.92 | 2.19 | (-11.21, -2.63) | **0.002** |
| Sensitivity analysis: further adjusted for additional clinical complications*† |  |  |  |  |
| Number of breastfeeding sessions | -0.33 | 0.08 | (-0.48, -0.18) | **<0.001** |
| Any breastfeeding | -5.60 | 1.96 | (-9.43, -1.77) | **0.004** |
| Any preoperative breastfeeding‡ | -6.94 | 1.76 | (-10.39, -3.49) | **<0.001** |

**Notes:**
Each line represents an individual model.
\*Variables accounted for by the energy balancing weighting process included birth weight-for-age z-score (WAZ), prenatal CCHD diagnosis, infant delivery mode, infant race, Hispanic ethnicity, insurance type, Child Opportunity Index 2.0 score, single ventricle physiology, aortic obstruction, major genetic syndrome, extracardiac anomaly, preoperative factors including mechanical circulatory support, shock, or neurological deficit, highest level of preoperative respiratory support, preoperative enteral feeding, prostaglandin E1 days, Society of Thoracic Surgeons–European Association for Cardio-Thoracic Surgery (STAT) mortality category, age at surgery, weight at surgery, sternum left open postoperatively, postoperative extracorporeal membrane oxygenation (ECMO). Human milk volume refers to the base diet during the neonatal period, not including fortification.
†Sensitivity analyses further adjusted for fixed effects of cardiac arrest, diaphragm dysfunction, chylothorax, duration of postop mechanical ventilation, and weight trajectory (weight-for-age z-score change, birth to discharge).
‡Infants with no preoperative feeding were excluded from these models.
Abbreviations: CI = confidence interval, OR = odds ratio, SE = standard error

Each additional BF session during the neonatal period was associated with around half a day shorter length of stay (0.31–0.65 shorter, p<0.001). Similarly, neonates with any BF had an estimated 8.65 days shorter LOS (p<0.001), and any preoperative BF was associated with 6.92 days shorter LOS (p=0.002). Both human milk and BF findings were consistent in sensitivity analyses with further adjustment for direct effects of multiple clinical complications (Table 5), in addition to the 20 key covariates already accounted for by the energy balancing weighting process (see Detailed Methods). In subgroup analyses of patients with single ventricle physiology, 100% human milk as the neonatal base diet, compared to 0%, was associated with 16.81 days shorter LOS (4.15–29.47 days shorter p=0.009), and each additional BF session was associated with 1.16 days shorter LOS (0.33–2.00 days shorter, p=0.006).

## Discussion

In this large, multisite study with detailed neonatal nutrition data, we found that increased human milk percentage during the neonatal period was not significantly associated with NEC, sepsis, or other infectious complications, but was strongly associated with shorter LOS. We also found that initiation of bovine-based fortification or formula was associated with subsequent NEC diagnosis, even among infants with established human milk feeding.

### Human milk percentage and NEC

Few CCHD studies to date have incorporated detailed, daily nutrition data to examine dose-dependent relationships between human milk and NEC. Although our finding of no significant association between human milk percentage and NEC differs from preterm literature, it aligns with a single center case-control study in CCHD by Christian et al..^43^ Evaluating nutrition in the 7 days preceding NEC diagnosis, they found that infants with NEC had lower human milk intake and higher exposure to bovine-derived fortification, though neither association reached statistical significance (p=0.10). The authors speculated that a small number of NEC cases (2.8% prevalence) may have limited their ability to detect an effect. Although the NEC prevalence in our study was similar (3.0%), we did not observe a comparable trend of lower human milk percentage in the NEC group. Interestingly, the human milk percentages in Christian et al.’s study (24.6% NEC cases, 45.9% matched controls) were far lower than in our multisite cohort (79.3%, 75.6%). It is possible that the relatively high rate of human milk feeding throughout the neonatal period in our cohort could have contributed to a NEC stage II–III prevalence that was lower than previous reports (eg, 3.9%–7.1% in a recent meta-analysis).^44^ Additionally, our inclusion criteria requiring at least 7 days of neonatal enteral feeding potentially excluded highly unstable infants and early, severe NEC cases that limited feeding or resulted in death before cardiac surgery. Therefore, our findings of no association between human milk percentage and NEC may not be fully generalizable to all infants with CCHD.

### Fortification or formula initiation and NEC

We identified a relationship between a new exposure to bovine-derived fortification or formula and subsequent development of NEC. This result suggests that, for some infants, exposure to even a small amount of bovine-derived formula via fortification or the base diet could contribute to the development of NEC. The results were consistent in multiple sensitivity analyses including adjustment for maximum pre-NEC enteral volume. Furthermore, the fortification level (kcal/oz) did not differ between NEC cases and controls. This finding is an important contribution to the CCHD nutrition literature, although replication in independent cohorts is needed to confirm the association and further evaluate its potential causal relevance. While fortification can be helpful in mitigating growth challenges related to heart failure and the operative course, there is currently little evidence to inform CCHD fortification practices.

Our results align with previous single-site investigations of preterm infants.^37,38,45^ In one study of preterm, human-milk fed infants who required surgery for NEC,^38^ 51% developed surgical NEC within 7 days of their first exposure to bovine-derived fortification or formula, including several infants who had tolerated human milk for more than a month before fortification or formula initiation. In another study, the likelihood of developing NEC was 12.32 times greater on the 4th day following powdered human milk fortifier initiation.^37^ It is possible that the higher osmolality of fortified feeding could play a role, however a recent systematic review noted no consistent evidence linking higher osmolality to adverse gastrointestinal events including NEC.^46^ There are, to our knowledge, no studies investigating nutrition osmolality specifically in CCHD populations.

Although there is extensive preclinical literature outlining various human milk bioactive components that confer protection against NEC,^47^ the mechanisms by which fortification or formula exposure could contribute to NEC development are unclear. Hypotheses include bovine protein intolerance,^48^ changes to the gut microbiome,^48^ differences in lipid digestion and absorption,^49^ and formula-related differences in intestinal hypoxia and splanchnic oxygenation,^50,51^ but the limited investigation to date has focused on preterm populations, is controversial,^52^ and may or may not be applicable in CCHD. Our findings highlight the critical need for risk stratification tools to identify infants with CCHD who may not tolerate bovine-derived fortification or formula introduction, particularly considering recent NEC-related lawsuits involving formula exposure.^52^ Predictive biomarkers are emerging and have potential to guide practice, but current results remain preliminary.^21,53–56^

A related unanswered question is whether the type of nutrition fortification could play a role in the development of NEC. Three recent studies have explored alternative fortification types.^19,57^ First, in a clinical trial of human milk-derived fortifier (Prolacta) compared to bovine-derived fortifier, Blanco et al. reported 12% lower rates of all stages of NEC, although this study was not powered for the rarer outcome of stage II–III NEC.^19^ Additionally, the base diet was not consistent between the intervention and control groups (eg, infants in the control group could consume 0–100% human milk), raising questions of whether the observed effects were due to the human-milk derived fortifier, lower exposure to bovine-derived formula as the base diet, or both. Second, Woodgate et al. examined NEC rates following an institutional practice change to use extensively hydrolyzed formula for feed fortification, compared to polymeric formula pre-implementation.^57^ While the overall rate of all stages of NEC did not change, the authors noted a lower rate of “severe” NEC (stage III) in the post-implementation group. However, this finding was not tested for statistical significance, no potential confounders were considered, and the rates of human milk feeding and fortification differed between groups. Finally, Palm et al. identified no difference in fortification or formula type between infants with stage I (benign hematochezia) vs. stages II–III NEC, but excluded infants with no NEC-related symptoms.^58^ While our results were unchanged when adjusting our analysis to consider fortification type (broadly defined as “standard” or “specialized”), our sample was not powered to explore the effect of hydrolyzed formula fortification on NEC, and no infants in the matched groups received human milk-derived fortifier. Thus, future work is needed to clarify optimal fortification practices in CCHD.

### Length of stay

We found that both increased human milk percentage and higher BF frequency during the neonatal period were strongly associated with shorter hospital LOS. These results align with a large, national single ventricle registry study, which reported that infants with greater human milk and BF exposure had shorter LOS for both stage 1 and stage 2 palliation hospitalizations in multiple propensity score-matched analyses.^20^ As this previous investigation also found strong associations between human milk/BF and reduced NEC, sepsis, and infection, the authors speculated that some of the LOS benefit could have been mediated by reductions in these diseases. Our findings contradict this hypothesis, although direct BF was relatively rare in this cohort and these low BF rates introduce some uncertainty. It is possible that human milk/BF may have lowered non-NEC feeding intolerance or gastrointestinal distress^59^ in our sample, with an associated reduction in LOS, but we did not measure general feeding intolerance and this seems unlikely to fully explain the large effect size.

Investigations of human milk/BF and LOS are often subject to reverse causality, in which prolonged hospitalization can negatively impact lactation. To address this, nutrition exposures in this study were limited to the neonatal period, which is a relatively short time to maintain human milk and BF, limiting the likelihood of this bias. It is also possible that only the “healthiest” infants, who would inherently have a shorter LOS, were encouraged to BF. However, in addition to the use of rigorous causal inference methods to reduce bias by balancing more than 20 variables that reflect an infant’s clinical course, we further adjusted for the direct effects of additional indicators of clinical instability that contribute to longer hospitalizations, and results were consistent. Limited preclinical evidence indicates that human milk may drive neonatal cardiomyocyte metabolism^60^ and improve wound healing,^61^ highlighting potential avenues for future research, but mechanisms to explain our results are currently unknown.

### Limitations

While our large, multisite study used methods for causal inference to address limitations of previous literature, we acknowledge that the retrospective nature prevents full confidence in the causal direction of our results, and residual confounding cannot be excluded. Disease risk score matching accounts for numerous relevant variables; however, the case-control analysis was limited by the necessary assignment of a counterfactual diagnosis age to control groups to maintain the causal pathway. Additionally, we did not have data on maternal factors known to impact lactation (eg, maternal intention to BF, pumping frequency, diabetes),^62^ which are likely relevant. As we collected detailed neonatal nutrition data (plus 2 days before discharge and NEC), we did not have full nutrition data for infants diagnosed with outcomes beyond 30 days. However, we conducted sensitivity analyses including only infants diagnosed with NEC by 30 days old, and results were unchanged. Human milk volumes from BF were estimated, which could affect the results. However, BF did not occur frequently in our sample, and the addition of BF volumes had limited impact on most infants’ human milk percentage. Donor human milk use could impact the results; however donor human milk use was limited in our sample (2.9% of the neonatal volume). EHR data is subject to differences in documentation. Similarly, diagnostic criteria for outcomes could have varied across centers, although detailed data definitions, derived from the PC4 registry whenever possible, were employed to minimize this variation. Our study design excluded some infants that may be prone to clinical instability and adverse outcomes including NEC, such as premature infants, those requiring ECMO before surgery, and patients with limited enteral feeding exposure. Therefore, the findings may not generalize to the highest-risk CCHD populations, premature infants with CHD, and those patients requiring prolonged preoperative support. Finally, although CoRe-PCICS sites vary widely in size and location, participating sites may differ from non-participating sites; thus, our results may not fully generalize.

### Conclusion

In this multisite cohort, the percentage of human milk received during the neonatal period was not associated with reduced rates of NEC, sepsis, or infection. There was, however, a relationship between bovine-derived fortification or formula initiation and an increased likelihood of NEC, even in cases where human milk feeding was well established. Additionally, increased neonatal human milk feeding and direct BF were strongly associated with a shorter hospital LOS, although mechanisms to explain this finding are unclear. Future research is critically needed to determine optimal fortification strategies (type, timing, advancement, duration) for high-risk infants with CCHD, and to develop risk stratification tools to identify infants who may not tolerate bovine-derived fortification or formula introduction. In the absence of strong evidence to guide CCHD clinical nutrition practice, a cautious approach to bovine-derived fortification or formula introduction in infants at high risk for NEC may be warranted.

## Supporting information

Detailed Methods

## Data Availability

The analytical methods used in this study can be made available to researchers outside the CoRe-PCICS collaborative if requested. The multisite data are governed under the conditions of the data use agreements between participating institutions and the study coordinating center and will not be publicly available.

## Non-standard Abbreviations and Acronyms

BF: Breastfeeding
CCHD: Critical congenital heart disease
CoRe-PCICS: Collaborative Research from the Pediatric Cardiac Intensive Care Society
CPB: Cardiopulmonary bypass
LOS: Length of stay
NEC: Necrotizing enterocolitis
VVR: vasoactive-ventilation-renal

## Acknowledgments

We thank Julian Wolfson (PhD, Professor, Division of Biostatistics & Health Data Science, School of Public Health, University of Minnesota) for his consultation and guidance. We also thank Lina Thiel, Alexis Steiber, and Nellie Munn Swanson (graduate students, University of Minnesota) for their assistance with data cleaning. We would like to sincerely thank all participating CoRe-PCICS centers, research coordinators, and clinical staff for their invaluable contributions to this study.

## Supplemental Material

Detailed Methods

Tables S1–S5

Figures S1–S4

