## Supplementary material for "Human milk feeding, fortification initiation, and clinical outcomes in neonates with critical congenital heart disease: A multi-institutional study": Detailed Methods

**Supplemental Materials**

*Detailed methods*

**1. Outcomes information:** Outcomes were defined as per the Pediatric Cardiac Critical Care Consortium (PC4) manual, as follows.

Necrotizing enterocolitis (NEC) was defined as an acute reduction in the supply of oxygenated blood to the small intestine or large intestine, typically resulting in acidosis, abdominal distention, pneumatosis, and/or intestinal perforation, that prompts initiation of antibiotics or exploratory laparotomy. NEC was defined as cases meeting criteria for modified Bell's Stage II or III:

- Stage IIA (Definite, Mildly Ill)
    - Systemic signs: Temperature instability, apnea, bradycardia
    - Intestinal signs: Elevated pregavage residuals, mild abdominal distention, gross blood in stool, absent bowel sounds, abdominal tenderness
    - Radiographic signs: Ileus and pneumatosis intestinalis
    - Treatment: NPO; antibiotics for 7–10 days
- Stage IIB (Definite, Moderately Ill)
    - Systemic signs: Same as Stage IIA, plus mild metabolic acidosis and thrombocytopenia
    - Intestinal signs: Same as Stage IIA, plus abdominal cellulitis, and right lower quadrant mass
    - Radiographic signs: Same as Stage IIA, plus portal venous gas with or without ascites
    - Treatment: NPO; antibiotics for 14 days
- Stage IIIA (Advanced, Severely Ill, Bowel Intact)
    - Systemic signs: Same as Stage IIB, plus hypotension, bradycardia, respiratory acidosis, metabolic acidosis, disseminated intravascular coagulation, and neutropenia
    - Intestinal signs: Same as Stage II, plus signs of generalized peritonitis and marked abdominal tenderness and distention
    - Radiographic signs: Same as Stage IIB, plus definite ascites
    - Treatment: NPO; antibiotics for 14 days; fluid resuscitation; inotropic support; ventilator therapy; paracentesis
- Stage IIIB (Advanced, Severely Ill, Bowel Perforated)
    - Systemic signs: Same as Stage IIIA
    - Intestinal signs: Same as Stage IIIA
    - Radiographic signs: Same as Stage IIB, plus pneumoperitoneum
    - Treatment: Same as Stage IIA, plus surgery

Sepsis: Sepsis was defined as temperature instability and abnormal white blood cells (leukopenia or leukocytosis) and either (1) initiation or escalation of inotropic support or (2) initiation or escalation of invasive mechanical ventilation. In addition the patient must have been treated with antibiotics for >6 days.

<u>Other infectious complication</u>: The composite "other infectious complication" outcome included superficial and deep surgical site infections (SSI), central line-associated bloodstream infection (CLABSI), urinary tract infection (UTI), ventilator-associated pneumonia (VAP), non-VAP pneumonia, and preoperative respiratory virus. Definitions for each are as follows:

- Superficial SSI: Defined according to CDC criteria. The infection must occur during the cardiac intensive care unit (CICU) encounter or within 48 hours of CICU discharge. Only newly acquired superficial SSIs adjudicated by local infection control are included.
- Deep SSI: Defined according to CDC criteria and includes deep incisional infections and organ-space infections (e.g., mediastinitis). The event must occur during the CICU stay or within 48 hours of discharge. Only newly acquired infections adjudicated by local infection control are included.
- CLABSI: Defined according to CDC criteria. Events must occur during the CICU stay or within 48 hours of discharge and must be adjudicated by local infection control as newly acquired.
- UTI: Defined according to CDC criteria and includes catheter-associated and non–catheter-associated UTIs. Events must occur during the CICU stay or within 48 hours of discharge. Catheter-associated UTIs (CAUTIs) must be adjudicated by local infection control. Non-CAUTI UTIs do not require adjudication.
- Ventilator-associated pneumonia (VAP): Defined according to CDC criteria and must occur during the CICU encounter or within 48 hours of discharge. VAP events attributable to the CICU must be adjudicated by local infection control.
- Non-VAP pneumonia: Defined according to CDC criteria and includes any pneumonia occurring during the CICU encounter that does not meet VAP criteria.
- Preoperative viral respiratory infection: Defined as any documented viral respiratory infection diagnosed at any time during the hospitalization before surgery, based on clinical assessment or PCR testing.

**2. Exposure information:**

<u>Direct breastfeeding (BF) volume</u>: As very few BF sessions included volume measurement (e.g., from pre-post weights), we estimated maternal human milk volumes from BF based on published research documenting average BF milk transfer in term and hospitalized infants,[1,2] as follows:

- Day of life 0: each BF session was estimated to be 6 mL
- Day 1: 8 mL
- Day 2: 9 mL
- Day 3: 11 mL
- Day 4: 12 mL
- Day 5–7: 13 mL
- Day 8–10: 15 mL
- Day 11–14: 20 mL

- Day 15–17: 25 mL
- Day 18–20: 30 mL
- Day 21 and older: 35 mL

These numbers are intentionally conservative, to reflect the often fragile medical state of newborns with CCHD during the perioperative time.

**3. Statistical analysis**

Details of the disease risk score matching approach: For the case-control analyses, we used logistic regression to create disease risk scores for matching with the *MatchIt* package in R. Twenty variables with potential to impact the exposure and the outcome were used to create the disease risk scores:

- Birth weight-for-age z-score (WAZ)
- Prenatal (vs. postnatal) CCHD diagnosis
- Infant delivery mode (eg, vaginal, caesarean)
- Infant race
- Hispanic ethnicity
- Insurance type
- Child Opportunity Index 2.0 score
- Single ventricle physiology (yes/no)
- Aortic obstruction (yes/no)
- Major genetic syndrome (yes/no)
- Extracardiac anomaly (yes/no)
- Preoperative factors including mechanical circulatory support, shock, or neurological deficit (see further definitions at the end of this section)
- Highest level of preoperative respiratory support
- Any preoperative enteral feeding (yes/no)
- Number of days receiving prostaglandin E1
- Society of Thoracic Surgeons-European Association for Cardio-Thoracic Surgery (STAT) mortality category
- Age at surgery (days)
- Weight at surgery (kg)
- Sternum left open postoperatively (yes/no)
- Postoperative extracorporeal membrane oxygenation (ECMO) requirement (yes/no)

All variables except infant delivery mode, Child Opportunity Index, number of prostaglandin E1 days, and STAT category were defined per the PC4 manual:

- Pediatric Cardiac Critical Care Consortium. Data Definitions Manual v3.0. Accessed January 12, 2024. https://pc4.arbormetrix.com/Registry/static/pc4/html/datacollection.html

Matching strategy

Cases were matched to up to 10 controls with no replacement, using nearest neighbor matching and a caliper of 0.3 standard deviations of the logit to improve the quality of matches. For NEC, sepsis, and infection outcomes, no controls went unmatched. Details include:

| **Outcome: NEC** | **Case** | **Control** |
|---|---|---|
| All | 22 | 797 |
| Matched (effective sample size) | 22 | 194.09 |
| Matched (unweighted) | 22 | 198 |
| Unmatched (n, %) | 0 (0%) | 599 (75.2%) |
| **Outcome: Sepsis** | **Case** | **Control** |
| All | 31 | 783 |
| Matched (effective sample size) | 31 | 145.15 |
| Matched (unweighted) | 31 | 234 |
| Unmatched (n, %) | 0 (0%) | 549 (70.1%) |
| **Outcome: Other infectious complication** | **Case** | **Control** |
| All | 93 | 725 |
| Matched (effective sample size) | 92 | 382.12 |
| Matched (unweighted) | 92 | 406 |
| Unmatched (n, %) | 1 (1.08%) | 319 (44.0%) |

Details of the energy balancing weighting approach: For the length of stay (LOS) analyses, we used the *WeightIt* package in R to calculate weights via energy balancing as described by Huling et al.,[3] which reduces bias and supports causal inference for continuous outcomes. The same variables listed above for disease risk score matching were used to create weights for human milk and BF exposures.

The weighting strategy for analysis of LOS as a function of human milk percentage resulted in a well-balanced cohort with a coefficient of variation of 0.43, mean absolute deviation of 0.33, and entropy of 0.09. The weighted effective sample size was 641.1 compared with an unweighted sample size of 758, indicating that the procedure achieved covariate balance while retaining most of the available information for subsequent outcome modeling. For analysis of LOS as a function of BF, the coefficient of variation was 0.42, mean absolute deviation was 0.28, and entropy 0.10. The weighted effective sample size was 643.98.

Weights were not trimmed or truncated. For the human milk percentage analysis, weights ranged from 0.025 to 2.85, with one participant receiving a very small weight. For the any-breastfeeding analysis, weights ranged from 0.014 to 4.20, with 44 participants (5.8%) receiving very small weights. The weighted effective sample sizes were 641.1 and 644.0, respectively, compared with an unweighted sample size of 758. Full weight distributions are shown in Figure S1.

Covariate balance for all analyses can be seen in Figure S2 and Table S2.

Missing data: There were no missing data in the full sample. For variables that were unknown or undocumented in the electronic health record, an "Unknown" category was used and included in the matching or weighting approaches. Affected covariates were Hispanic ethnicity (n=60 unknown) and birth delivery mode (n=12 unknown).

Adjustment for site: We considered center-level effects in our analysis plan. However, because there were only 22 NEC cases distributed across 25 sites, the sparse number of events per center limited the reliable estimation of center effects in the NEC models. We therefore excluded site from the NEC models. Importantly, sensitivity analyses that included site produced essentially unchanged estimates and interpretation.

For example, for fortification initiated within 5 days of NEC diagnosis (Table 4), the odds ratio was 3.10 (p=0.025) without site adjustment and 3.14 (p=0.025) with site added as a random effect. Similarly, for the percentage of human milk received (Table 3), results were nearly identical with and without site adjustment for NEC (OR=1.35, p=0.656 vs. OR=1.34, p=0.663), sepsis (OR=0.58, p=0.297 vs. OR=0.60, p=0.370), and infection (OR=1.53, p=0.229 vs. OR=1.61, p=0.211).

The length-of-stay analysis was also unchanged: human milk feeding was associated with a 9.33-day shorter length of stay (p<0.001) without site adjustment and a 9.01-day shorter length of stay (p<0.001) with site added as a fixed effect.

Thus, although site was not included in the primary models because of the sparse distribution of events across centers, sensitivity analyses demonstrate that inclusion of site does not materially alter the magnitude, direction, or statistical significance of the findings.

PC4 definitions for key covariates:

1. Shock: Root definition

Shock is defined as "a state of inadequate tissue perfusion". A modern definition according to Simeone states that shock is a "clinical condition characterized by signs and symptoms which arise when the cardiac output is insufficient to fill the arterial tree with blood under sufficient pressure to provide organs and tissues with adequate blood flow." A historic definition according to Blalock in 1940 is that "Shock is a peripheral circulatory failure, resulting from a discrepancy in the size of the vascular bed and the volume of the intravascular fluid".

- Simeone FA. Shock, trauma and the surgeon. Ann Surg. 1963;158(5):759-774. doi:10.1097/00000658-196311000-00004
- Blalock A. Principles of surgical care: shock and other problems. St. Louis, MO: C.V. Mosby; 1940.

*a. Shock, Persistent at time of surgery*

Code this factor if the patient had a metabolic acidosis with pH < 7.2 and/or Lactate > 4 mmol / liter at the time of OR Entry Date and Time.

*b. Shock, Resolved at time of surgery*

Code this factor if the patient had a metabolic acidosis with pH < 7.2 and/or Lactate > 4 mmol / liter at any time after the date and time of admission to the hospital but not at the time of OR Entry Date and Time. This factor should be coded if shock was present at any time after the date and time of admission to the hospital but not at the time of OR Entry Date and Time, including situations where shock was present after admission to the hospital where this operation

2. Preoperative/Preprocedural mechanical circulatory support (IABP, VAD, ECMO, or CPS)

Code this factor if the patient is supported with mechanical support, of any type (IABP, VAD, ECMO, or CPS), for resuscitation/CPR or support, at the time of OR Entry Date and Time.

3. Preoperative neurological deficit

Code this factor if the patient has any deficit of neurologic function identified by the care team (during the hospitalization of this operation prior to the time of OR Entry Date and Time).

*Supplemental figures and figure legends*

**Figure S1.** Distribution of matching and balancing weights across analyses

**a.** NEC outcome

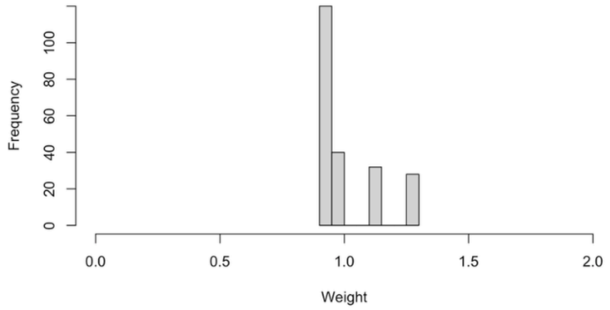

**b.** Sepsis outcome

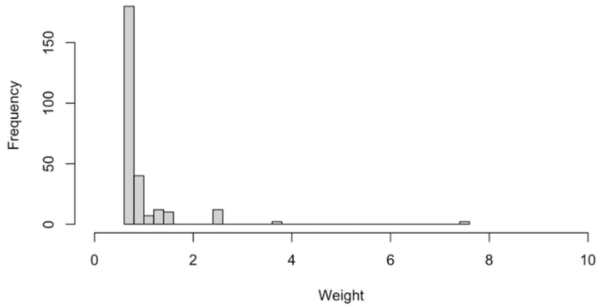

**c.** Composite other infection outcome

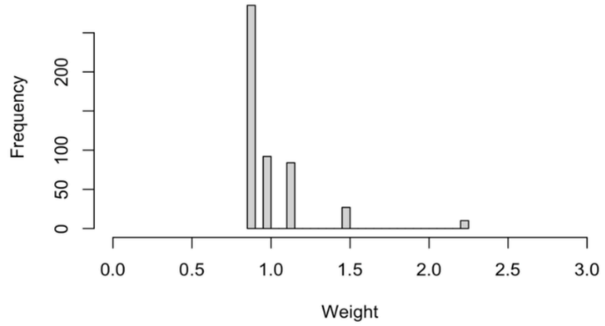

**d.** Length of stay outcome (human milk percentage)

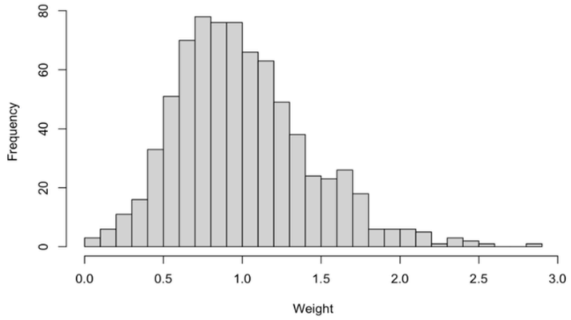

**e.** Length of stay outcome (any direct breastfeeding)

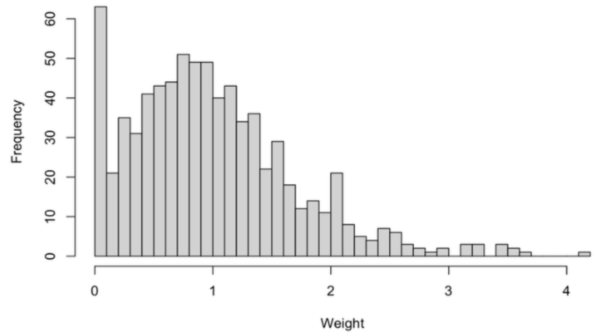

**Figure S2.** Covariate balance for disease risk score matched or energy balancing weighted analytical cohorts

**a.** NEC outcome

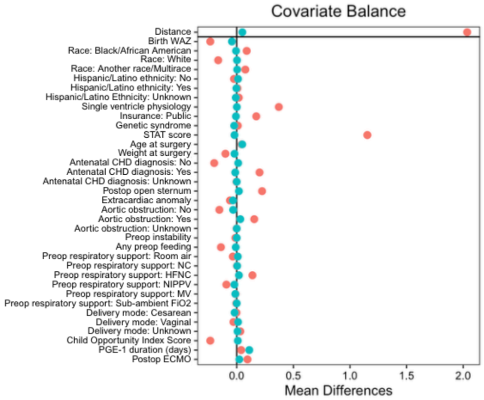

**b.** Sepsis outcome

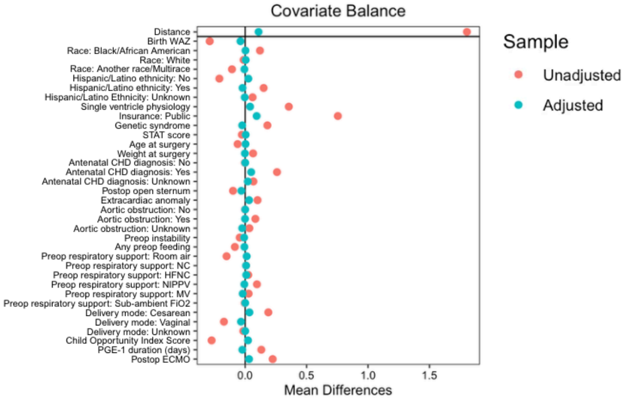

**c.** Composite other infection outcome

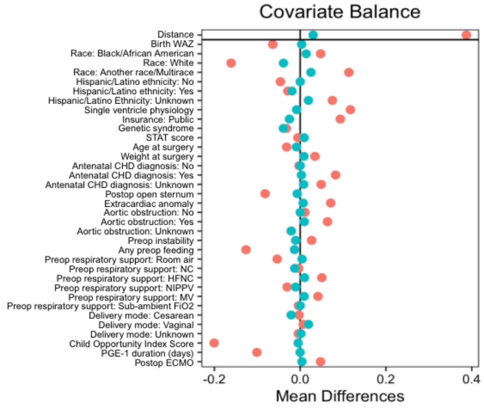

**d.** Length of stay outcome (human milk percentage)

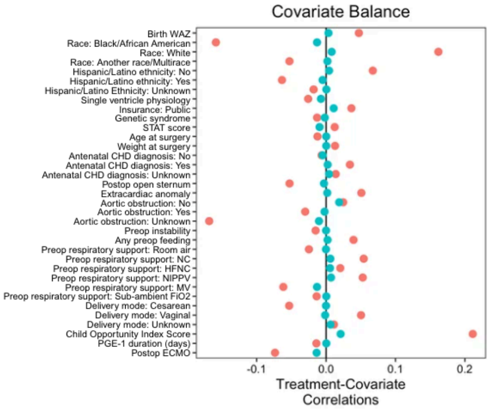

**e.** Length of stay outcome (any direct breastfeeding)

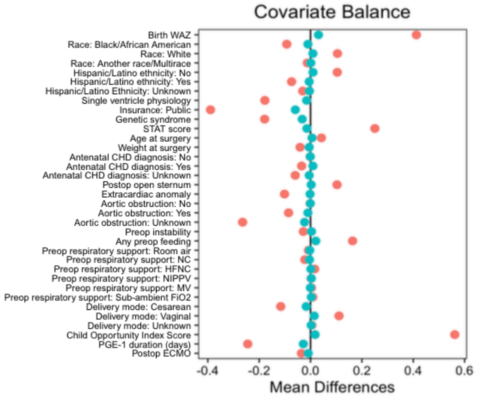

Note: These plots represent covariate balance for matching (NEC, sepsis, infection) or weighting (length of stay) using human milk percentage and any breastfeeding exposures.

**Figure S3.** Distribution of neonatal human milk percentage in disease risk score matched cohorts with and without NEC

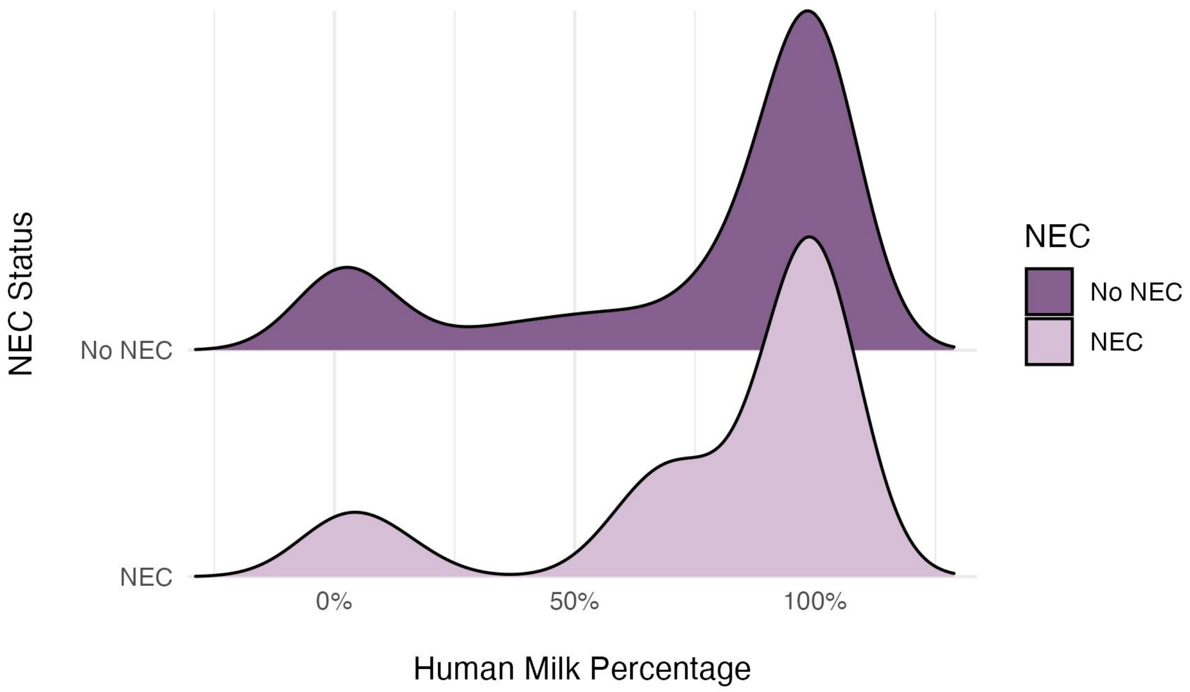

**Figure S4.** Unadjusted associations between neonatal breastfeeding, human milk exposure, and hospital length of stay. (A) Length of stay by any neonatal breastfeeding; boxes show median and interquartile range, points are individual infants. (B, C) Length of stay by number of neonatal breastfeeding episodes and by percentage of neonatal enteral volume as human milk; lines are ordinary least-squares fits with 95% confidence bands. Panels display unweighted, unadjusted data; the fitted lines are descriptive and do not incorporate the energy-balancing weights or covariate adjustment used in the primary analysis.

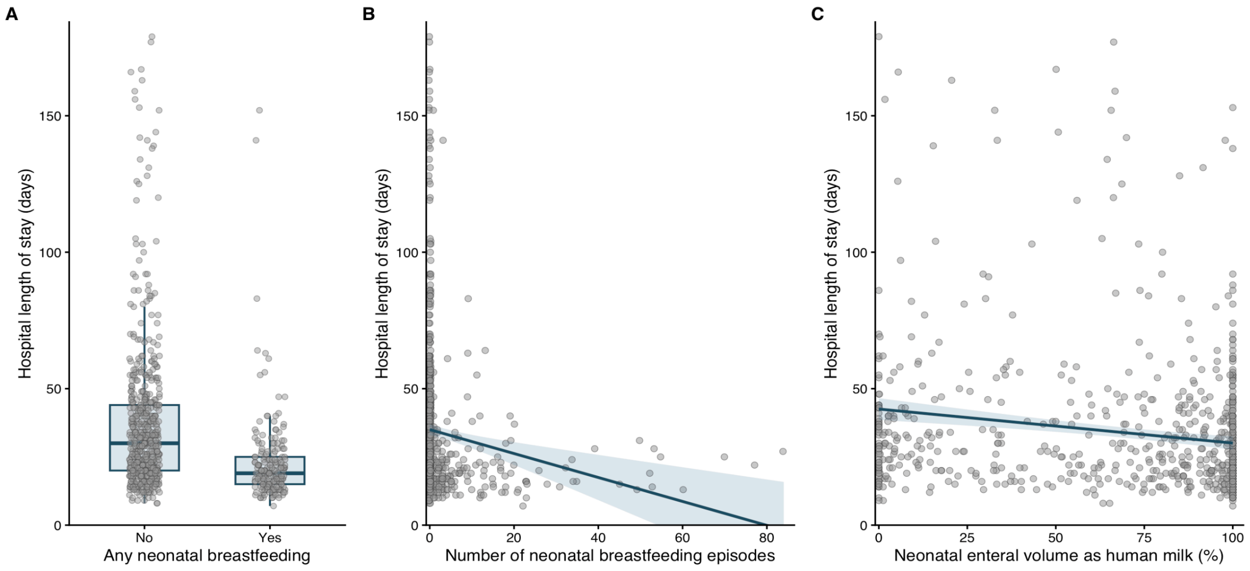

*Supplemental tables and supporting information*

**Table S1. Participating CoRe-PCICS Institutions**

| |
|---|
| Advocate Children's Hospital |
| Ann & Robert H Lurie Children’s Hospital of Chicago |
| Boston Children's Hospital |
| Children's Healthcare of Atlanta |
| Children's Hospital of Alabama |
| Children's Hospital of Illinois at OSF Healthcare |
| Children's Hospital of Michigan |
| Children's National Hospital |
| Cincinnati Children's Hospital Medical Center |
| Cohen Children's Medical Center of New York |
| Duke University Hospital |
| Johns Hopkins Hospital |
| Le Bonheur Children's Hospital |
| Lucile Packard Children's Hospital Stanford |
| Medical University of South Carolina Children's Hospital |
| Nationwide Children's Hospital |
| Norton Children’s Hospital |
| New York Presbyterian - Morgan Stanley Children's Hospital |
| Primary Children's Hospital |
| Rady Children's Hospital San Diego |
| Riley Hospital for Children at Indiana University Health |
| Seattle Children's Hospital |
| Texas Children's Hospital |
| UF Health Shands Children's Hospital |
| University of Minnesota M Health Fairview Masonic Children’s Hospital |

**Table S2. Covariate balance before and after matching or weighting, across five exposure–outcome analyses**

| | NEC | | Sepsis | | Infectious complication | | Length of stay: Human milk % | | Length of stay: Any breastfeeding | |
|---|---|---|---|---|---|---|---|---|---|---|
| | *Matched: SMD* | | *Matched: SMD* | | *Matched: SMD* | | *Energy weighted: Correlation* | | *Energy weighted: SMD* | |
| **Covariate** | **Unadjusted** | **Adjusted** | **Unadjusted** | **Adjusted** | **Unadjusted** | **Adjusted** | **Unadjusted** | **Adjusted** | **Unadjusted** | **Adjusted** |
| Disease risk score | 2.03 | 0.05 | 1.80 | **0.11** | 0.39 | 0.03 | — | — | — | — |
| Birth weight-for-age z-score | -0.23 | -0.04 | -0.29 | -0.04 | -0.06 | 0.00 | 0.05 | 0.00 | 0.41 | 0.03 |
| **Race** | | | | | | | | | | |
| Black/African American | 0.21 | -0.01 | 0.27 | 0.01 | 0.12 | 0.04 | -0.16 | -0.01 | -0.09 | -0.01 |
| White | -0.33 | 0.00 | -0.03 | 0.01 | -0.32 | -0.08 | 0.16 | 0.01 | 0.10 | 0.01 |
| Another race or multirace | 0.17 | 0.01 | -0.36 | -0.02 | 0.25 | 0.05 | -0.05 | 0.00 | -0.01 | 0.00 |
| **Ethnicity** | | | | | | | | | | |
| Not Hispanic/Latino | -0.05 | 0.03 | -0.42 | 0.05 | -0.10 | 0.00 | 0.07 | 0.00 | 0.10 | 0.01 |
| Hispanic/Latino | 0.02 | -0.01 | 0.32 | -0.05 | -0.08 | -0.05 | -0.06 | -0.01 | -0.07 | -0.01 |
| Unknown | 0.06 | -0.03 | 0.18 | -0.01 | 0.22 | 0.06 | -0.02 | 0.00 | -0.03 | 0.00 |
| Single-ventricle physiology | 0.77 | 0.00 | 0.73 | 0.08 | 0.24 | -0.02 | -0.03 | -0.01 | -0.18 | -0.02 |
| STAT score | 1.15 | -0.02 | 0.75 | 0.09 | 0.09 | -0.03 | 0.04 | 0.01 | -0.39 | -0.06 |
| Age at surgery | 0.04 | 0.05 | 0.18 | -0.02 | -0.03 | -0.04 | -0.01 | 0.00 | -0.18 | -0.03 |
| Weight at surgery | -0.10 | -0.02 | -0.03 | 0.00 | -0.01 | 0.01 | 0.01 | -0.01 | 0.25 | -0.01 |
| **Antenatal diagnosis of CHD** | | | | | | | | | | |
| No | -0.69 | 0.05 | -0.15 | 0.01 | -0.07 | -0.02 | -0.01 | 0.00 | 0.04 | 0.01 |
| Yes | 0.70 | -0.05 | 0.15 | -0.01 | 0.08 | 0.02 | 0.01 | 0.00 | -0.04 | -0.01 |
| Unknown | -0.05 | 0.00 | -0.05 | 0.00 | -0.06 | 0.00 | -0.01 | 0.00 | 0.00 | 0.00 |
| Sternum left open | 0.48 | 0.04 | 0.57 | **0.11** | 0.17 | 0.01 | 0.03 | 0.00 | -0.03 | 0.01 |
| Extracardiac abnormality | -0.17 | -0.10 | 0.15 | 0.05 | 0.12 | 0.02 | 0.01 | 0.00 | -0.06 | -0.01 |
| **Aortic obstruction** | | | | | | | | | | |
| No | -0.32 | -0.07 | -0.20 | -0.06 | -0.16 | -0.01 | -0.05 | 0.00 | 0.10 | 0.00 |
| Yes | 0.32 | 0.07 | 0.20 | 0.06 | 0.14 | 0.01 | 0.05 | 0.00 | -0.10 | 0.00 |
| Unknown | -0.04 | 0.00 | -0.04 | 0.00 | 0.10 | 0.00 | 0.02 | 0.02 | 0.00 | 0.00 |
| Preoperative instability | -0.03 | 0.00 | 0.20 | 0.00 | 0.16 | 0.02 | -0.03 | 0.00 | -0.09 | -0.01 |
| Public insurance | 0.37 | -0.02 | 0.07 | -0.05 | -0.04 | -0.04 | -0.17 | -0.01 | -0.26 | -0.02 |
| Genetic syndrome | 0.03 | -0.06 | -0.13 | -0.03 | 0.07 | -0.03 | -0.02 | 0.00 | -0.03 | 0.00 |
| Preoperative feeding | -0.28 | -0.02 | -0.18 | -0.01 | -0.26 | -0.03 | 0.04 | 0.00 | 0.16 | 0.02 |
| **Preoperative respiratory support** | | | | | | | | | | |
| Room air | -0.08 | 0.02 | -0.41 | 0.03 | -0.12 | 0.01 | -0.02 | 0.00 | -0.01 | 0.00 |

| | | | | | | | | | | |
|---|---|---|---|---|---|---|---|---|---|---|
| Nasal canula | 0.01 | 0.01 | 0.02 | 0.02 | -0.01 | -0.05 | 0.05 | 0.01 | -0.02 | 0.00 |
| High flow nasal canula | 0.31 | 0.04 | 0.07 | 0.02 | 0.13 | 0.03 | 0.02 | 0.01 | 0.02 | 0.00 |
| Non-invasive positive pressure ventilation | -0.44 | -0.10 | 0.23 | -0.02 | -0.10 | -0.03 | 0.05 | 0.01 | 0.00 | 0.00 |
| Mechanical ventilation | -0.02 | -0.03 | 0.06 | -0.04 | 0.09 | 0.02 | -0.06 | -0.01 | 0.00 | 0.00 |
| Sub-ambient FiO2 | -0.06 | 0.00 | -0.06 | 0.00 | -0.07 | 0.00 | -0.01 | 0.00 | 0.01 | 0.00 |
| **Mode of delivery** | | | | | | | | | | |
| Cesarean | -0.01 | -0.04 | 0.38 | 0.07 | -0.01 | -0.04 | -0.05 | 0.00 | -0.12 | -0.02 |
| Vaginal | -0.06 | 0.02 | -0.35 | -0.07 | 0.01 | 0.04 | 0.05 | 0.00 | 0.11 | 0.01 |
| Unknown | 0.15 | 0.03 | -0.13 | 0.00 | -0.04 | 0.02 | 0.01 | 0.01 | 0.01 | 0.00 |
| Child Opportunity Index | -0.23 | 0.01 | -0.27 | 0.02 | -0.20 | 0.00 | 0.21 | 0.02 | 0.56 | 0.02 |
| Prostaglandin days | 0.04 | **0.11** | 0.13 | -0.02 | -0.10 | 0.00 | -0.01 | 0.00 | -0.24 | -0.03 |
| Postoperative ECMO | 0.28 | 0.07 | 0.51 | 0.07 | 0.17 | 0.01 | -0.07 | -0.01 | -0.04 | -0.01 |
| *Covariates with value > 0.10 (n)* | *19* | *1* | *26* | *1* | *18* | *0* | *4* | *0* | *15* | *0* |

Abbreviations: CHD, congenital heart disease; DRS, disease risk score; ECMO, extracorporeal membrane oxygenation; LOS, length of stay; SMD, standardized mean difference; STAT, The Society of Thoracic Surgeons–European Association for Cardio-Thoracic Surgery

**Table S3.** Type of initial fortification before the matched necrotizing enterocolitis age

| Fortification type | Full case-control cohort (n = 220) | NEC (n = 22) | No NEC (n = 198) |
|---|---|---|---|
| Standard term formula | 35 (15.9) | 4 (18.2) | 31 (15.6) |
| Partially hydrolyzed | 32 (14.5) | 2 (9.1) | 30 (15.2) |
| Extensively hydrolyzed | 21 (9.5) | 2 (9.1) | 19 (9.6) |
| Amino acid-based | 13 (5.9) | 1 (4.5) | 12 (6.1) |
| Human milk fortifier (bovine-derived) | 12 (5.5) | 1 (4.5) | 11 (5.6) |
| Other specialty (energy-dense, high-MCT) | 6 (2.8) | 4 (18.1) | 2 (1.0) |
| Preterm/post-discharge formula | 4 (1.8) | 0 (0.0) | 4 (2.0) |
| Not specified | 4 (1.8) | 0 (0.0) | 4 (2.0) |
| Not fortified before matched NEC age* | 93 (42.3) | 8 (36.4) | 85 (42.9) |

Notes: Data are n (%). Percentages are calculated within column among all 220 infants. Exposure is defined as the first fortification or formula received before the necrotizing enterocolitis diagnosis age of the matched case.
*No fortification or formula was received before the diagnosis age of the matched case. Infants in this category may have been fortified subsequently.

**Table S4.** Characteristics of the disease risk score matched cohort for the sepsis outcome

| | Full case-control cohort[a] | Sepsis (cases) | No sepsis (controls) | p value |
|---|---|---|---|---|
| | (N = 265) | (N = 31) | (N = 234) | |
| Sex at birth | | | | 0.902 |
| Male | 176 (66.4) | 20 (64.5) | 156 (66.7) | |
| Female | 88 (33.2) | 11 (35.5) | 77 (32.9) | |
| Race | | | | 0.921 |
| White | 176 (66.4) | 20 (64.5) | 156 (66.7) | |
| Black/African American | 61 (23.0) | 8 (25.8) | 53 (22.6) | |
| Another race or multirace | 28 (10.6) | 3 (9.7) | 25 (10.7) | |
| Ethnicity | | | | 0.933 |
| Hispanic/Latino | 87 (32.8) | 10 (32.3) | 77 (32.9) | |
| Not Hispanic/Latino | 149 (56.2) | 17 (54.8) | 132 (56.4) | |
| Not documented | 29 (10.9) | 4 (12.9) | 25 (10.7) | |
| Insurance status | | | | >0.999 |
| Public | 148 (55.8) | 17 (54.8) | 131 (56.0) | |
| Non-public or other | 117 (44.2) | 14 (45.2) | 103 (44.0) | |
| Child Opportunity Index | 42 (20, 68) | 40 (20, 66) | 43 (20, 68) | 0.883 |
| Prenatal CHD diagnosis | 199 (75.1) | 24 (77.4) | 175 (74.8) | 0.922 |
| Birth WAZ | -0.32 (1.10) | -0.44 (0.87) | -0.30 (1.13) | 0.519 |
| Single ventricle physiology | 133 (50.2) | 19 (61.3) | 114 (48.7) | 0.261 |
| Genetic syndrome | 32 (12.1) | 4 (12.9) | 28 (12.0) | >0.999 |
| Extracardiac abnormality | 53 (20.0) | 8 (25.8) | 45 (19.2) | 0.534 |
| Any preoperative feeding | 177 (66.8) | 20 (64.5) | 157 (67.1) | 0.933 |
| Surgery age (days) | 6 (4, 11) | 6 (3, 11) | 6 (5, 10) | 0.478 |
| Weight (kg) at surgery | 3.32 (0.54) | 3.30 (0.47) | 3.32 (0.55) | 0.783 |
| STAT score 4 or 5 | 184 (69.4) | 25 (80.6) | 159 (67.9) | 0.217 |
| Sternum left open | 164 (61.9) | 22 (71.0) | 142 (60.7) | 0.362 |
| Postoperative ECMO | 31 (11.7) | 8 (25.8) | 23 (9.8) | 0.021 |
| Sepsis | 31 (11.7) | – | – | |
| Sepsis diagnosis age (days) | 21 (10, 36) | – | – | – |
| % of volume as HM (base diet) before sepsis diagnosis age | 75.46 (35.28) | 70.55 (40.77) | 76.13 (34.51) | 0.410 |
| Exclusive HM (base diet) before sepsis diagnosis age | 120 (46.7) | 17 (54.8) | 103 (45.6) | 0.437 |
| Any breastfeeding before sepsis | 36 (13.6) | 1 (3.2) | 35 (15.0) | 0.130 |
| Length of stay (days) | 34 (22, 53) | 75 (44, 103) | 33 (21, 47) | <0.001 |

Notes:
a. Descriptive statistics are n (%) for categorical variables, mean (SD) for continuous variables except median (25%, 75%) for Child Opportunity Index, age at surgery, sepsis diagnosis age, and hospital length of stay.
Abbreviations: CHD = congenital heart disease, ECMO = extracorporeal membrane oxygenation, HM = human milk, STAT = Society of Thoracic Surgeons-European Association for Cardio-Thoracic Surgery, WAZ = weight-for-age z-score

**Table S5.** Characteristics of the disease risk score matched cohort for the “other infectious complication” outcome[a]

| | Full case-control cohort[b] | Infection (cases) | No infection (controls) | p value |
|---|---|---|---|---|
| | (N=498) | (N=93) | (N=405) | |
| Sex at birth | | | | 0.764 |
| Male | 317 (63.7) | 57 (61.3) | 260 (64.2) | |
| Female | 180 (36.1) | 36 (38.7) | 144 (35.6) | |
| Race | | | | 0.372 |
| White | 289 (58.0) | 48 (51.6) | 241 (59.5) | |
| Black/African American | 77 (15.5) | 16 (17.2) | 61 (15.1) | |
| Another race or multirace | 132 (26.5) | 29 (31.2) | 103 (25.4) | |
| Ethnicity | | | | 0.253 |
| Hispanic/Latino | 86 (17.3) | 14 (15.1) | 72 (17.8) | |
| Not Hispanic/Latino | 360 (72.3) | 65 (69.9) | 295 (72.8) | |
| Not documented | 52 (10.4) | 14 (15.1) | 38 (9.4) | |
| Insurance status | | | | 0.830 |
| Public | 254 (51.0) | 46 (49.5) | 208 (51.4) | |
| Non-public or other | 244 (49.0) | 47 (50.5) | 197 (48.6) | |
| Child Opportunity Index | 46 (24, 72) | 43 (22, 66) | 47 (24, 72) | 0.526 |
| Prenatal CHD diagnosis | 364 (73.1) | 69 (74.2) | 295 (72.8) | 0.892 |
| Birth WAZ | −0.25 (1.08) | −0.27 (1.12) | −0.25 (1.08) | 0.865 |
| Single ventricle physiology | 182 (36.5) | 36 (38.7) | 146 (36.0) | 0.718 |
| Genetic syndrome | 100 (20.1) | 18 (19.4) | 82 (20.2) | 0.960 |
| Extracardiac abnormality | 112 (22.5) | 22 (23.7) | 90 (22.2) | 0.872 |
| Any preoperative feeding | 320 (64.3) | 57 (61.3) | 263 (64.9) | 0.588 |
| Surgery age (days) | 6 (4, 9) | 5 (4, 8) | 6 (4, 9) | 0.581 |
| Weight (kg) at surgery | 3.30 (0.53) | 3.30 (0.54) | 3.30 (0.52) | 0.926 |
| STAT score 4 or 5 | 273 (54.8) | 51 (54.8) | 222 (54.8) | >0.999 |
| Sternum left open | 260 (52.2) | 50 (53.8) | 210 (51.9) | 0.828 |
| Postoperative ECMO | 32 (6.4) | 9 (9.7) | 23 (5.7) | 0.237 |
| Other infectious complication | 93 (18.7) | – | – | – |
| Infection diagnosis age (days) | 16 (12, 24) | – | – | – |
| % of volume as HM (base diet) before infection diagnosis age[c] | 76.1 (35.5) | 80.2 (33.9) | 75.1 (35.8) | 0.200 |
| Exclusive HM (base diet) before infection diagnosis age[c] | 251 (52.6) | 53 (57.6) | 198 (51.4) | 0.286 |
| Any breastfeeding before infection diagnosis age | 91 (18.3) | 13 (14.3) | 79 (19.4) | 0.255 |
| Length of stay (days) | 29 (20, 44) | 40 (26, 62) | 27 (19, 41) | <0.001 |

Notes:

a. Infectious complications are defined as surgical site infection, central line-associated blood stream infection, urinary tract infection, pneumonia, and preoperative respiratory viral infection.

b. Descriptive statistics are n (%) for categorical variables, mean (SD) for continuous variables except median (25%, 75%) for Child Opportunity Index, age at surgery, and hospital length of stay.

c. Defined only among cases; n = 92 with non-missing exclusive human milk status.

Abbreviations: CHD = congenital heart disease, ECMO = extracorporeal membrane oxygenation, HM = human milk, STAT = Society of Thoracic Surgeons-European Association for Cardio-Thoracic Surgery, WAZ = weight-for-age z-score

**Table S6.** Primary cardiac diagnoses of infants who developed NEC within 5 days of first exposure to bovine-derived formula or fortification (n=7)

| | Single ventricle | Aortic obstruction[a] |
|---|---|---|
| Unbalanced AV canal defect | Yes | Yes |
| IAA + VSD | No | Yes |
| HLHS (mitral atresia, aortic atresia) | Yes | Yes |
| HLHS | Yes | Yes |
| L-TGA | No | No |
| DILV, hypoplastic transverse aortic arch | Yes | Yes |
| HLHS | Yes | Yes |

Notes:
a. Aortic obstruction was defined per Parker and Landstrom, 2021 (doi: 10.1161/JAHA.120.019006) as "structural and stenotic lesions that block left ventricular filling, output, and systemic blood flow. Among these are hypoplastic left heart syndrome, aortic stenosis, bicuspid aortic valve, coarctation of the aorta, and interrupted aortic arch."
The prevalence of single ventricle physiology was n=6 (75%) in the NEC group with fortification/formula within 5 days, and 17 (68%) in the matched controls with formula/fortification within 5 days (p=0.708). The prevalence of aortic obstruction was 7 (88%) in the NEC group, and 18 (72%) in the controls (p=0.373).
Abbreviations: AV = atrioventricular, DILV = double inlet right ventricle, HLHS = hypoplastic left heart syndrome, IAA = interrupted aortic arch, L-TGA = levo-transposition of the great arteries, VSD = ventricular septal defect.

*Supplemental references*

1. Feldman-Winter L, Kellams A, Peter-Wohl S, et al. Evidence-based updates on the first week of exclusive breastfeeding among infants ≥35 weeks. *Pediatrics*. 2020;145(4):e20183696. doi:10.1542/peds.2018-3696

2. Santoro W, Martinez FE, Ricco RG, Jorge SM. Colostrum ingested during the first day of life by exclusively breastfed healthy newborn infants. *J Pediatr*. 2010;156(1):29-32. doi:10.1016/j.jpeds.2009.07.009

3. Huling JD, Greifer N, Chen G. Independence weights for causal inference with continuous treatments. *J Am Stat Assoc*. 2024;119(546):1657-1670. doi:10.1080/01621459.2023.2213485
